# Bayesian back-calculation and nowcasting for line list data during the COVID-19 pandemic

**DOI:** 10.1101/2020.12.08.20238154

**Authors:** Tenglong Li, Laura F. White

## Abstract

Surveillance is the key of controling the COVID-19 pandemic, and it typically suffers from reporting delays and thus can be misleading. Previous methods for adjusting reporting delays are not particularly appropriate for line list data, which usually have lots of missing values that are non-ignorable for modeling reporting delays. In this paper, we develop a Bayesian approach that dynamically integrates imputation and estimation for line list data. We show this Bayesian approach lead to accurate estimates of the epidemic curve and time-varying reproductive numbers and is robust to deviations from model assumptions. We apply the Bayesian approach to a COVID-19 line list data in Massachusetts and find the reproductive number estimates correspond more closely to the control measures than the ones based on the reported curve.

## 1 Introduction

Surveillance plays a pivotal role in controlling the COVID-19 pandemic and has been used to provide guidance for government responses to the pandemic [1, 2]. A prerequisite for effective surveillance is to have daily case counts that are ideally defined based on infection dates (called the incidence curve) or, at a minimum, symptom onset dates (called the epidemic curve), which are biologically meaningful [3, 4, 5]. However what is most frequently recorded are case reporting dates, which tend to be either the date when an infected individual was tested, tested positive, or reported to public health authorities. The processes that impact the timing of case reporting date, namely obtaining and reporting test results, vary due to a large number of factors, including individual healthcare seeking behaviors, testing availability, or other factors that are not related to disease pathogenesis [6, 7]. This means that the reported curve (daily counts based on case reporting dates) have artificial noise that blurs the underlying epidemiological signal best described by infection dates, or secondarily by symptom onset dates [8, 9]. It also means that it is challenging to obtain timely estimates of the reproductive number as the most recently reported cases likely represent infection events that occurred some time in the past [10]. As these reported curves are often used to estimate reproductive numbers for surveillance and determining the efficacy of interventions, it is important that these cases are reported as close to the actual infection dates as possible [11, 12].

Infection dates are the most epidemiologically meaningful dates as they directly inform infection events and the reproductive numbers. However, obtaining infection dates is very challenging because infection events are not directly observable [12]. This is especially the case for COVID-19 due to significant pre-symptomatic transmission [13]. Typically, infection dates can only be obtained based on a strong parametric assumption about the distribution of incubation period, which is challenging to estimate [12, 14, 15]. On the other hand, symptom onset dates are more readily observed and in many settings captured for a subset of cases [3]. While symptom onset dates are not as helpful as infection dates, they are still linked to the epidemiology of infectious disease and are typically more proximate to infection events than case reporting dates [3, 16]. This makes the epidemic curve more informative than the reported curve for estimating reproductive numbers [17]. In practice, the major barrier for getting the epidemic curve is that symptom onset dates are still missing for many cases. This makes imputation of reporting delays, which are defined as the lags between symptom onset dates and case reporting dates for individual cases [8, 9, 10], a prerequisite for estimating the epidemic curve. In this paper, we rely on line list data which contains individual case reporting dates and symptom onset dates for some to impute individual reporting delays for all individuals.

Based on observed and imputed reporting delays, there are two steps to recover the epidemic curve from the reported curve. The first step is back-calculation which requires one to back-calculate symptom onset date based on case reporting date for each case [3]. Therefore, the epidemic curve is estimated by the daily case counts based on symptom onset dates rather than case reporting dates. The second step is nowcasting, which is needed because of the reported curve is right truncated, i.e., any case that is reported after the final reporting date (but potentially has symptom onset before the final reporting date) is unavailable for analysis [5]. The consequence of this right truncation issue is that the back-calculated counts of cases that show symptoms on days close to the final reporting date are likely incomplete as some of those cases are actually reported after the final reporting date and unavailable for back-calculation [17]. Hence, nowcasting is the task of modeling and appropriately increasing those case counts. The idea of back-calculation and nowcasting is illustrated by Fig 1.

**Figure 1:**
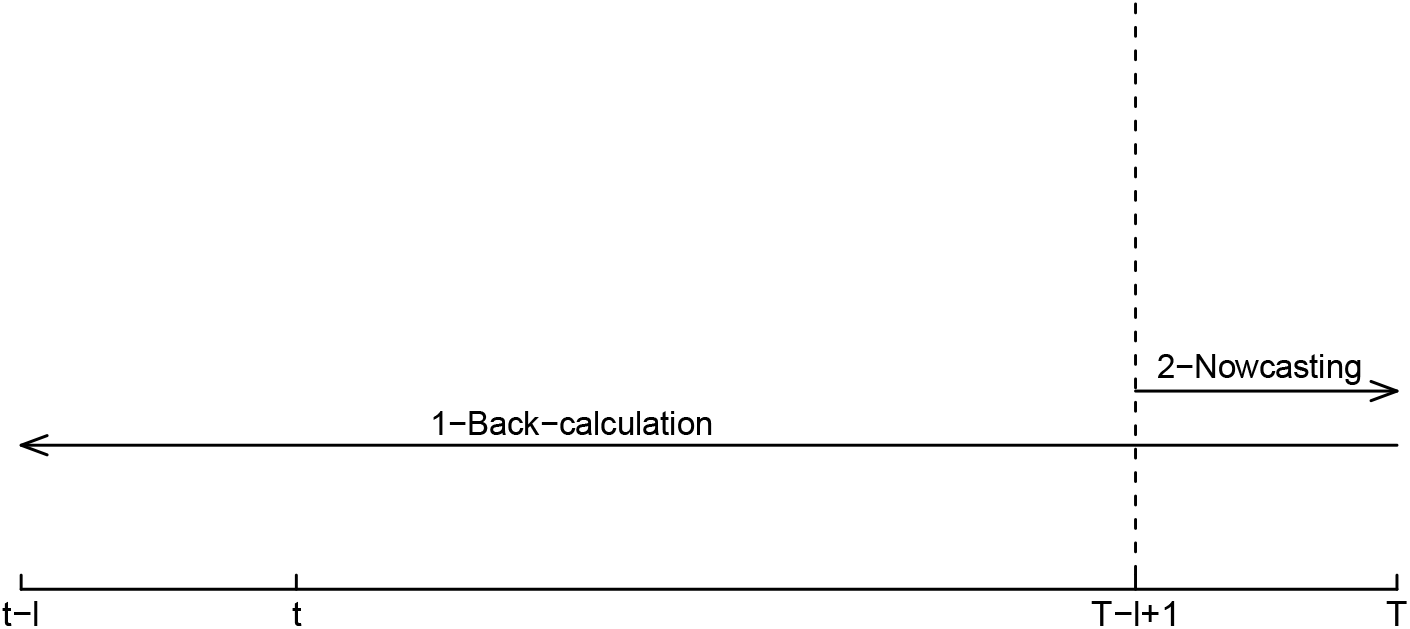
Illustration of back-calculation and nowcasting. Assuming *t* and *T* are the first and last reporting day in a line-list data, one needs to first back-calculate the daily case counts that cover the period from day *t* − *l* to day *T* based on reporting delays, where *l* is the maximum reporting delay. The next step is nowcasting, which is to upscale the back-calculated counts for the period from day *T* − *l* + 1 to day *T*.

Most previous work estimates the epidemic curve either by the one-step approach where one models the reporting delay distribution and/or case counts directly [5, 18], or by the two-step approach where one imputes missing reporting delays first (the imputation step) and then recovers the epidemic curve based on the imputed values (the estimation step) [3]. The reporting delay distribution is usually modeled based on the *reporting triangle*, a summary of the empirical distributions of reporting delays based on symptom onset dates. [5, 18, 19]. Since the *reporting triangle* does not take missing reporting delays into account, the one-step approach is based on observed reporting delays only and typically is time invariant. With such limitations, the two-step approach is generally preferred for a line list data where the missing reporting delays are non-ignorable. In this approach, the imputation step usually assumes symptom onset dates are missing at random conditional on case reporting dates and other available covariates in a line list data [3]. Usually, the imputed reporting delays in the two-step approach are not dynamically updated by the model of the reporting delay distribution, and they may be biased and have large variance. More importantly, making inference about the estimated epidemic curve would be difficult for the two-step approach since the variance associated with the imputation step is not taken into account by the estimation step.

In this paper, we develop a Bayesian framework that dynamically integrates the imputation step and the estimation step. Our Bayesian framework has five components: (1) inference of the reporting delay distribution based on case reporting dates; (2) imputation of missing reporting delays; (3) back-calculation; (4) nowcasting; and (5) reproductive number estimation using the *EpiEstim* method [20, 21]. The Bayesian framework is simple to implement and suitable for estimating the epidemic curve. We demonstrate the robustness of our framework by simulating an epidemic wave similar to the first COVID-19 outbreak under various conditions, such as changes in reporting delay distribution, violation of model assumptions, and incomplete surveillance data. We also demonstrate that the 95% Bayesian credible intervals have good coverage rate even under moderately undesirable conditions and therefore can lead to reliable inferences. We apply this Bayesian method to COVID-19 data in Massachusetts show that this method creates estimates of the epidemic curve and the reproductive number consistent with the COVID-19 dynamics in Massachusetts.

## 2 Materials and methods

### 2.1 Imputation of the missing reporting delays

For a line list data, we denote individual case reporting date and symptom onset date as *r*_*i*_ and *o*_*i*_, respectively, for individuals *i* = 1, …, *n*. Therefore, an individual reporting delay is defined as *d*_*i*_ = *r*_*i*_ − *o*_*i*_ and we assume *d*_*i*_ ∈ [0, *l*] for the missing *d*_*i*_. Moreover, we assume reporting starts from day 1 and ends at day *T* in the line list data. We use *t* to denote dates and *t* could be a negative integer. The maximum delay *l* can be decided based on the observed reporting delays as well as prior knowledge about the reporting system. The entire reporting period (from day 1 to day *T* in the line list data) can be thought of as the composition of consecutive small reporting periods, such that the reporting delay distribution is stable during each small reporting period. For example, for COVID-19 line list data we can define each week as the small reporting period under the assumption that the reporting delay distribution is unlikely to change sharply within each week. Then, we define *X*_1_ as the *n* × *p* matrix containing the indicators of the small reporting periods and *X*_2_ as the indicator of whether a case is reported on weekends, assuming there are *p* small reporting periods in total (for instance *p* is the number of weeks in the study period). The reporting delay distribution is then modeled for a single spatial region based on case reporting dates:

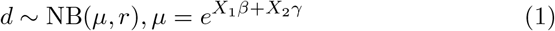

where *r* and *µ* are the size (dispersion) and mean parameters for negative binomial distribution.

Sometimes a reporting system improves over time and the reporting delays are significantly shortened after a specific date *t*_*c*_. In this case, Eq (1) is modified as:

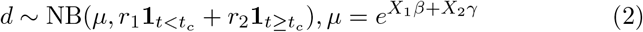

where **1**_*A*_ is the indicator of whether the condition *A* is met. In this formulation, Eq (2) has two dispersion parameters: *r*_1_ corresponds to dates prior to *t*_*c*_ and *r*_2_ corresponds to dates equal or later than *t*_*c*_. Based on Eq (1), the posterior distribution for imputing the missing reporting delays is:

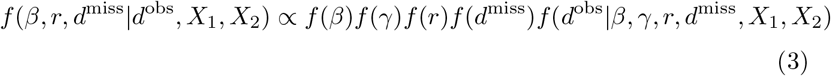

where *d*^miss^ represents all the missing *d*_*i*_ and *d*^obs^ represents all the observed *d*_*i*_. Using uninformative priors for *β, γ, r*, and *d*^miss^, imputation of *d*^miss^ is done via the following Gibbs sampler:

1. sample from *f* (*d*^miss^| *β, γ, r, X*_1_, *X*_2_)
2. sample from *f* (*β*| *γ, r, d*^miss^, *β, d*^obs^, *X*_1_, *X*_2_)
3. sample from *f* (*γ*| *r, d*^miss^, *β, d*^obs^, *X*_1_, *X*_2_)
4. sample from *f* (*r*| *d*^miss^, *β, γ, d*^obs^, *X*_1_, *X*_2_)

where *f* (*d*^miss^| *β, γ, r, X*_1_, *X*_2_) is a truncated negative binomial distribution whose upper bound is *l*. The above posterior distribution and Gibbs sampler are similarly defined for Eq (2).

In reality, the reporting delay distribution is most likely defined based on symptom onset dates rather than case reporting dates. Since symptom onset dates are the target of imputation, it is impossible to build a model conditional on them. By defining the small reporting periods and modeling the reporting delay distribution for each of these periods, we aim to estimate the reporting delay distribution within each of these periods and thus collectively approximate the underlying reporting delay distribution defined by symptom onset dates. Intrinsically, our approach is data mining rather than statistical modeling of the reporting delay distribution.

### 2.2 Estimation of the epidemic curve and reproductive numbers

Back-calculation is straightforward given the imputed *d*^miss^ and *d*^obs^. The back-calculated counts 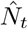, i.e., the case counts based on symptom onset dates in a line list data, is computed as:

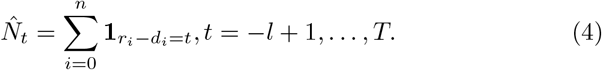

where 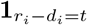 is the indicator of whether the *i*^*th*^ case showed symptoms on day *t*. Assuming the line list data includes all symptomatic cases, we can take 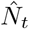 as the estimate of *N*_*t*_, the true number of cases who showed symptoms on day *t*, up to day *t* = *T* − *l*. Due to right-truncation 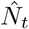 likely underestimates *N*_*t*_ for *t* = *T* − *l* + 1, …, *T*. To address this, we correct 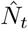 via a non-parametric nowcasting approach:

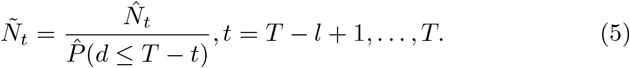

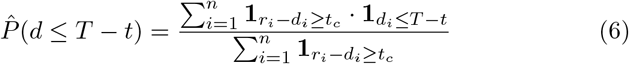

where *Ñ_t_* is the final estimate of *N*_*t*_ and 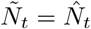 for *t* = −*l* + 1,…,*T* − *l*. If *t*_*c*_ is not provided, it implies there is no change in the reporting system and the indicator 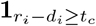 is always 1. In this case, 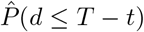 is the empirical cumulative density function of the line list data.

With the epidemic curve estimates *Ñ_t_,t* = −*l* + 1,…,*T*, the time-varying reproductive number estimates 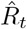 can be obtained based on *EpiEstim* [20, 21] *with a sliding window size τ*:

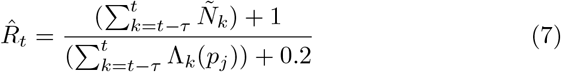

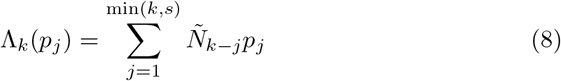

The serial interval distribution is needed for computing 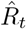: *s* is the maximum length of serial interval and *p*_*j*_ is the probability of a serial interval of *j* days. Since both the epidemic curve estimate *Ñ_t_* and reproductive number estimates 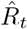 depend on the imputed reporting delays *d*^miss^, *Ñ_t_* and 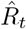 are computed based on the posterior sample of *d*^miss^ and updated by the Gibbs sampler for imputation, as well. Therefore, the final output of our Bayesian algorithm is a posterior sample of *Ñ_t_* and 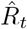. Statistical inference based on their Bayesian credible intervals incorporates the uncertainty about *d*^miss^.

### 2.3 Overview of simulation study

We simulated a local epidemic similar to COVID-19 using a branching process with the parameters based on COVID-19 literature [11, 14, 22, 23]. From this, we created a line list data based on the simulated epidemic wave (see details in appendix). By definition, the branching process started at day 1 and cases reported after day 60 were excluded in the line list data. For simulation scenarios, we vary three factors: data availability, the maximum reporting delay *l* assumption, and changes in the reporting delay distribution. We considered three possibilities regarding data availability: 1) complete data, 2) delayed surveillance initiation, and 3) real time estimation. The first scenario is ideal with the line list data covering the entire epidemic wave. In the second scenario, the line list data is only available after a certain date during the epidemic wave, possibly due to delays in initiating surveillance. In this case, we explored four different starting dates for the line list data to reflect various degrees to which earlier reports were lost and explore the impact of these delays on our approach. Third, we focus on estimation in the midst of the epidemic wave, which means the final reporting date in the line list data is prior to the end of the epidemic wave. We chose two different final reporting dates for the line list data: 1) before the peak of the reported curve, and 2) after the peak.

We also tested the case where we assumed *l* was 20 days for estimation when *l* actually was 25 days. We considered three possible scenarios regarding the changing dynamics of the reporting delay distribution over time. First, the reporting delay distribution remained unchanged and there was no improvement throughout the epidemic wave. The average reporting delay was 9 days in this case. Second, the reporting delay distribution sharply improved to an average of 4 days in the middle of the epidemic wave (*t*_*c*_ = day 30 based on symptom onset dates). Third, the reporting delay distribution was constantly and gradually improving during the epidemic wave. The average reporting delay gradually decreased from 9 days at the beginning to 4 days at the end of the epidemic wave.

We simulated 1000 line list datasets for each of the 18 different simulation scenarios. On average, the line-list data included about 5000 cases over 54 days. We randomly made the symptom onset dates missing for 60% of the cases, a percentage that was consistent with CDC line list data.

### 2.4 Line list data of COVID-19 cases in Massachusetts

We apply our method to a CDC line list data for Massachusetts with 85,627 COVID-19 cases. 823 cases were excluded from analysis due to negative reporting delays, which cannot be handled by our model. We excluded 5 cases with unusually large reporting delays reported before March 4, 2020. We set the maximum reporting delay to 60 days, marking 102 cases with longer reporting delays (ranging from 61 days to 117 days) as missing. Based on the data, these 102 reporting delays were clear outliers, potentially due to data entry errors. The final line list data contained 84,799 cases reported from March 4, 2020 to May 14, 2020 with symptom onset dates missing for 61.3% of the cases. Each of the 11 weeks was defined as the small reporting period for model estimation. The data and code are available at https://github.com/tenglongli/backandnow.

## 3 Results

### 3.1 Simulation results: complete line list data and delayed surveillance initiation

To ensure convergence of the Markov Chain Monte Carlo (MCMC) algorithm, the posterior sample was obtained based on 21,000 MCMC iterations with 1000 burn-in iterations for each of the 1000 simulated datasets. For all reproductive number estimation, the serial interval was assumed to follow the gamma distribution with the shape equal to 4.29 and the rate equal to 1.18 [14, 24], and the maximum serial interval was assumed to be 14 days. The median and 95% Bayesian credible intervals of the posterior samples of *Ñ_t_* and 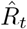 were extracted for each simulated dataset. The known epidemic curve and estimated reproductive numbers for each dataset served as the simulation benchmarks. To demonstrate the difference between the epidemic and reported curves, the reported curve and the reproductive number estimates based on it were also obtained for each dataset. The estimates were evaluated by two metrics: 1) the actual coverage rate of the 95% Bayesian credible interval based on 1000 simulated datasets, and 2) the root mean square error (RMSE) calculated as follows:

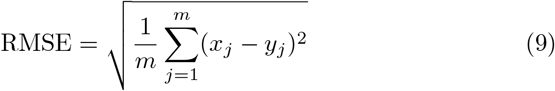

where *m* is the number of simulated datasets. *x*_*j*_ and *y*_*j*_ are the estimate and benchmark for *j*^*th*^ dataset.

As expected, the estimated epidemic curve and reproductive numbers were much closer to the simulation benchmark than the reported curve and the corresponding reproductive number estimates. With complete line list data, our model estimated true epidemic curve and the reproductive number well and was not sensitive to the changes in the reporting delay distribution (Fig 2). The estimates were not sensitive to the assumption about the maximum delay *l* across all simulation scenarios. For example, we illustrated the impact of the maximum delay assumption for complete line list data. (Fig 7, Fig 8 and Fig 9). Therefore, we only discuss the results obtained under the correct maximum delay assumption in the main text henceforth.

**Figure 2:**
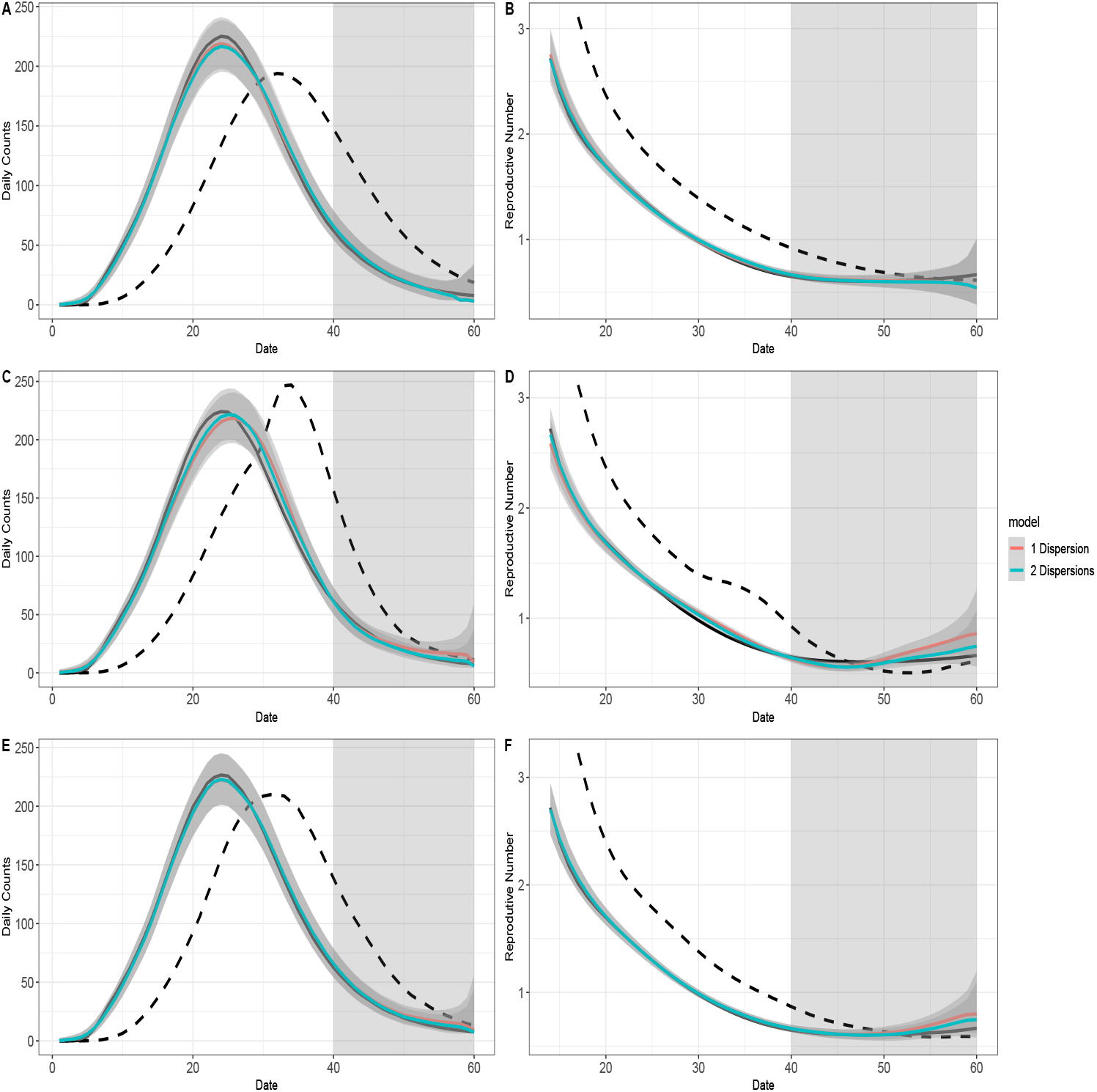
The model fit for complete data. For all graphs: the black solid curve corresponds to estimates based on the known epidemic curves and the black dashed curve corresponds to estimates based on the reported curves. The grey-shaded region superimposed on the curve depicts the 95% Bayesian credible interval and the grey-shade region on the right indicates the region of nowcasting. The colored curves represent different model choices. All values were averaged over 1000 simulated datasets with the correct *l*. A: The epidemic curve estimates if the reporting delay distribution was unchanged. B: The reproductive number estimates if the reporting delay distribution was unchanged. C: The epidemic curve estimates if the reporting delay distribution was sharply improved. D: The reproductive number estimates if the reporting delay distribution was sharply improved. E: The epidemic curve estimates if the reporting delay distribution was gradually improved. F: The reproductive number estimates if the reporting delay distribution was gradually improved.

**Figure 3:**
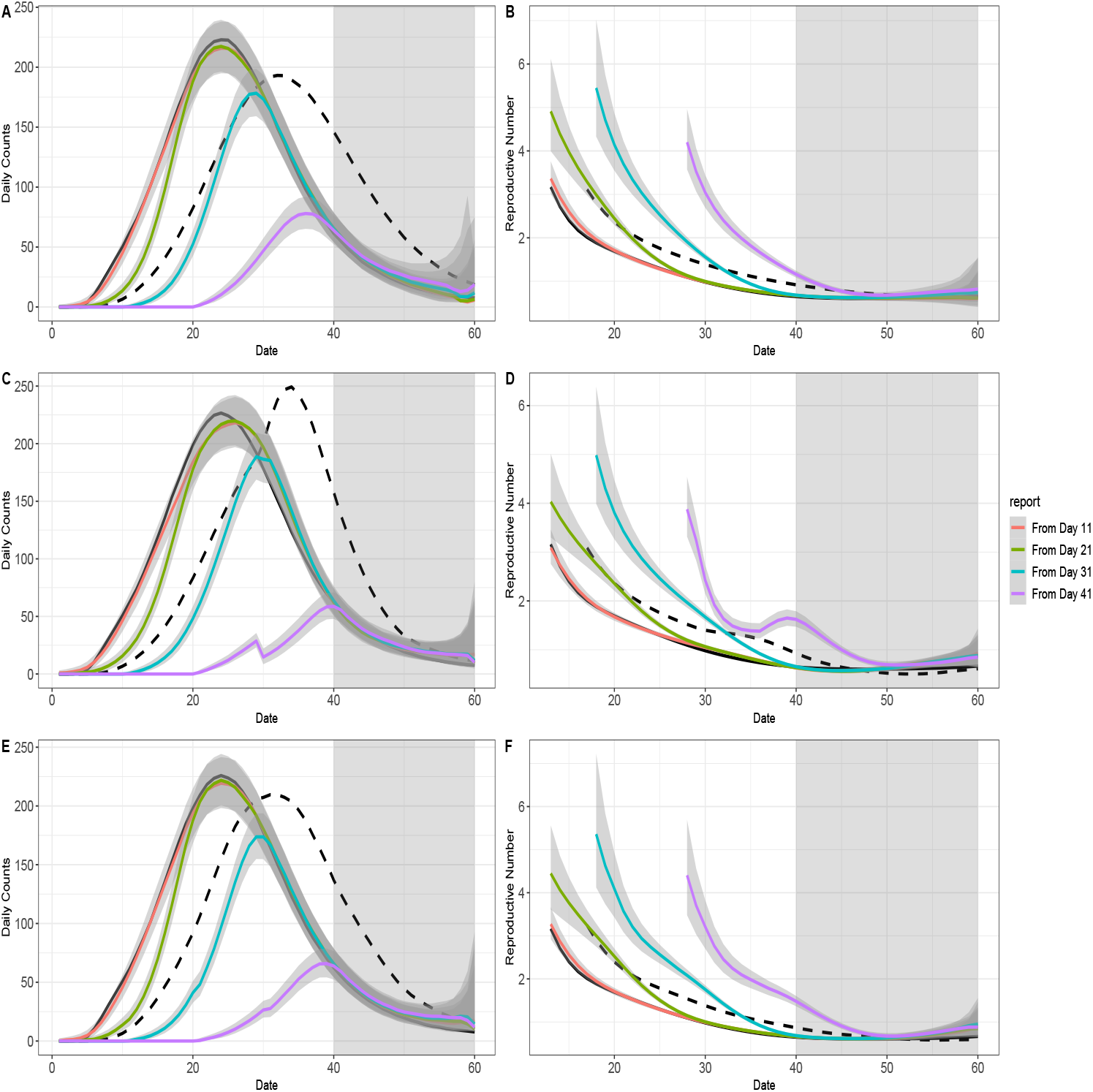
The model fit for data with no early report. For all graphs: the black solid curve corresponds to estimates based on the known epidemic curves and the black dashed curve corresponds to estimates based on the reported curves. The grey-shaded region superimposed on the curve depicts the 95% Bayesian credible interval and the grey-shade region on the right indicates the region of nowcasting. The colored curves represent different starting dates for the line-list data. All values were averaged over 1000 simulated datasets with the correct *l*. A: The epidemic curve estimates if the reporting delay distribution was unchanged. B: The reproductive number estimates if the reporting delay distribution was unchanged. C: The epidemic curve estimates if the reporting delay distribution was sharply improved. D: The reproductive number estimates if the reporting delay distribution was sharply improved. E: The epidemic curve estimates if the reporting delay distribution was gradually improved. F: The reproductive number estimates if the reporting delay distribution was gradually improved.

**Figure 4:**
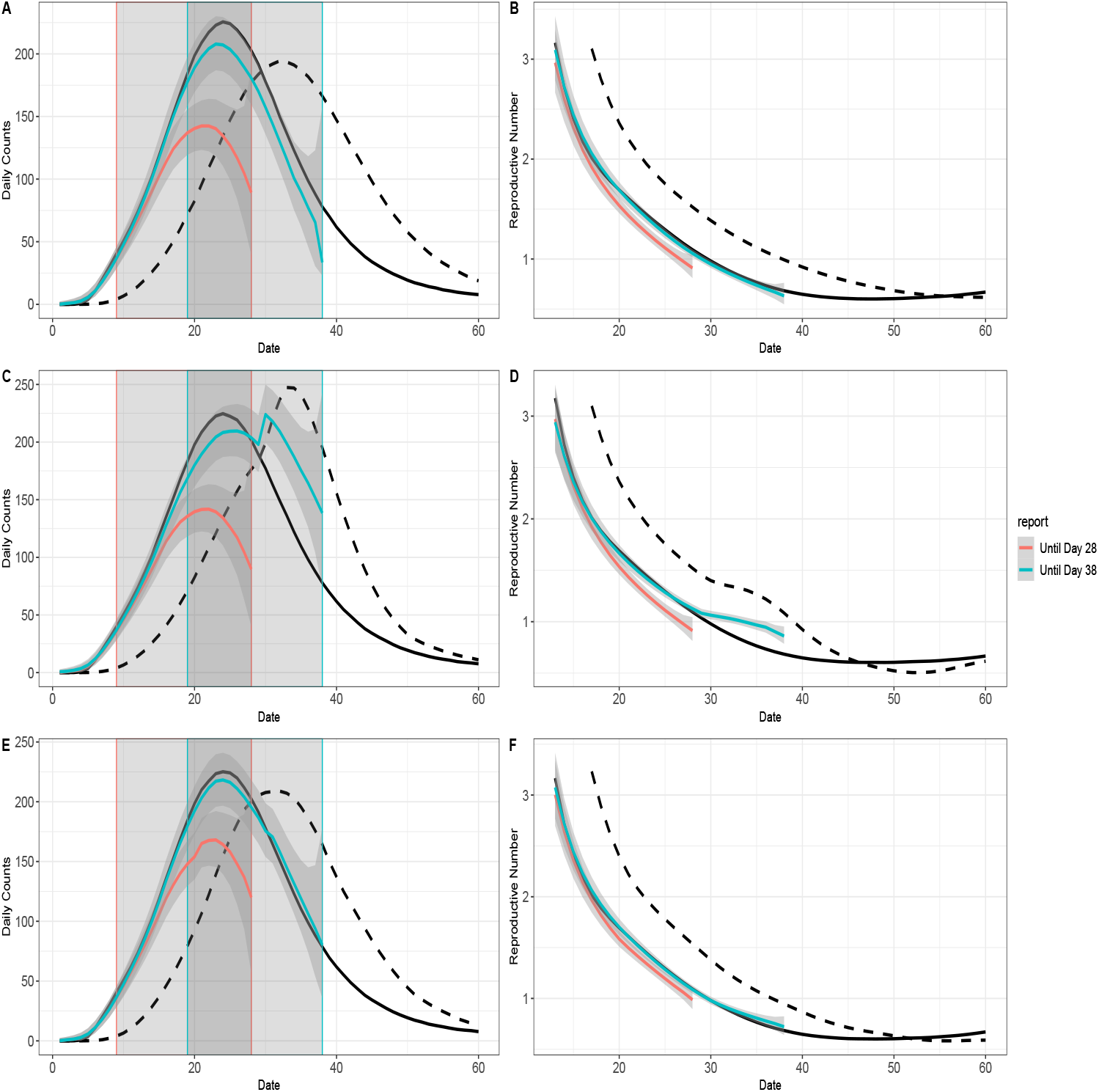
The model fit for an ongoing epidemic wave. For all graphs: the black solid curve corresponds to estimates based on the known epidemic curves and the black dashed curve corresponds to estimates based on the reported curves. The grey-shaded region superimposed on the curve depicts the 95% Bayesian credible interval. The colored curves represent different ending dates for line-list data, and their nowcasting regions are displayed as the gray-shaded areas with boundary lines in their corresponding colors. All values were averaged over 1000 simulated datasets. All values were averaged over 1000 simulated datasets with the correct *l*. A: The epidemic curve estimates if the reporting delay distribution was unchanged. B: The reproductive number estimates if the reporting delay distribution was unchanged. C: The epidemic curve estimates if the reporting delay distribution was sharply improved. D: The reproductive number estimates if the reporting delay distribution was sharply improved. E: The epidemic curve estimates if the reporting delay distribution was gradually improved. F: The reproductive number estimates if the reporting delay distribution was gradually improved.

**Figure 5:**
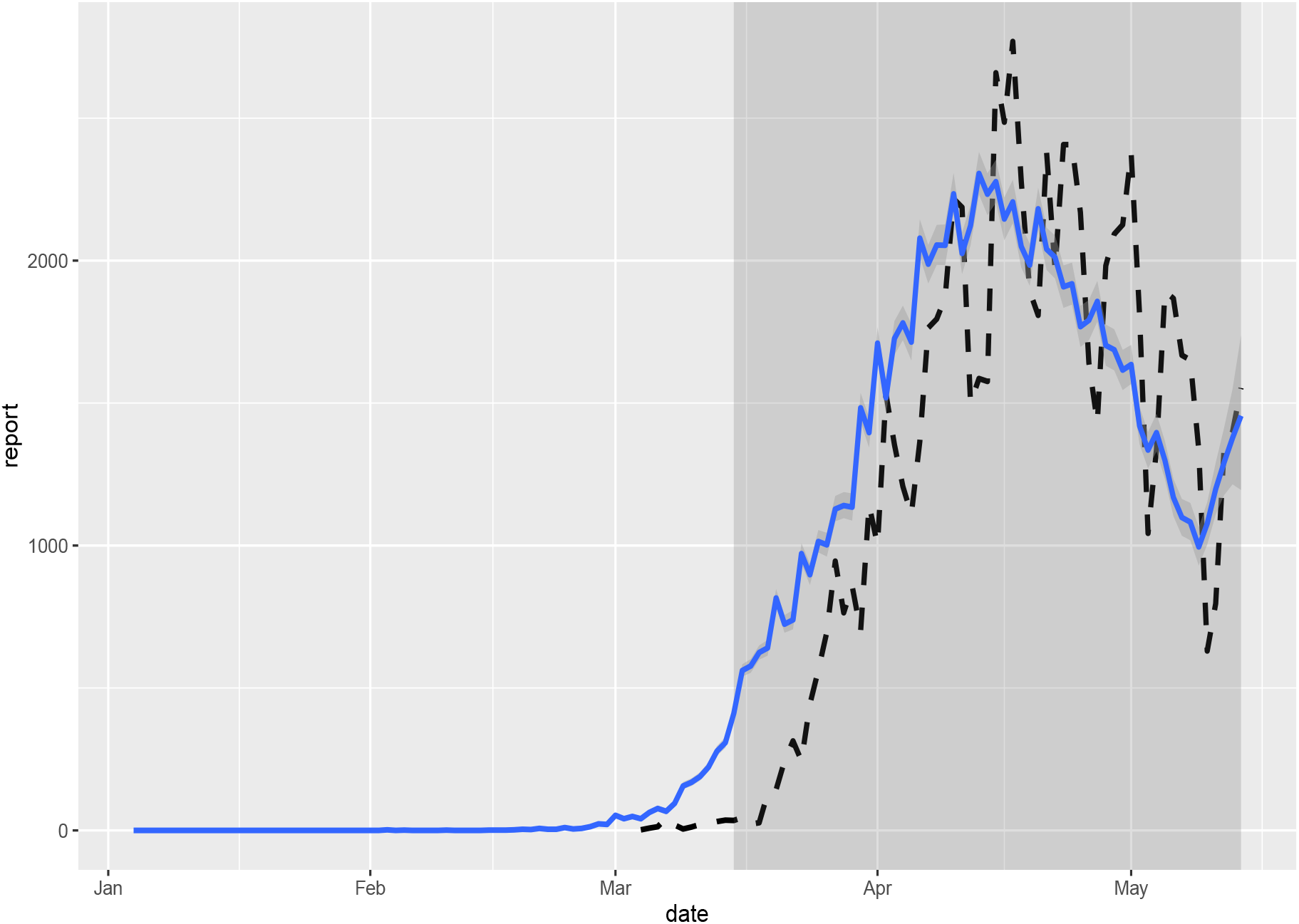
Estimated epidemic curve of COVID-19 in Massachusetts. The estimated epidemic curve was calculated based on weekly smoothing window and *l* = 60. The line-list data started on March 4, 2020 and ended on May 14, 2020. The earliest possible date that a case showed symptoms was February 1, 2020 and nowcasting started from March 16, 2020. The dashed curve represents the reported curve.

**Figure 6:**
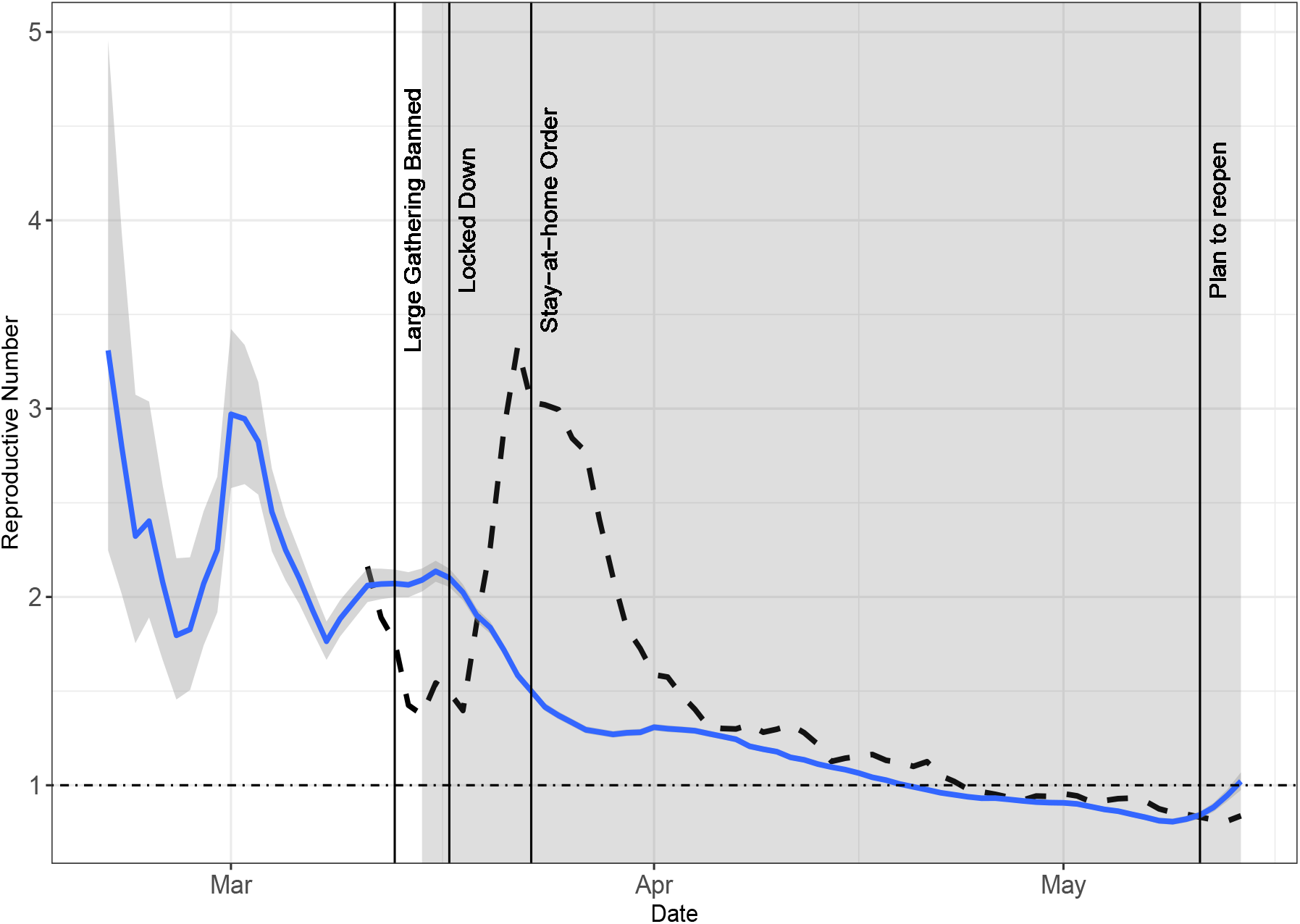
Estimated time-varying reproductive number of COVID-19 in Massachusetts. The estimates were calculated based on *EpiEstim* and a posterior sample of epidemic curve estimates. We identify the dates for four key policies: large gathering banned (March 13, 2020), lockdown (March 17, 2020), stay-at-home order (March 23, 2020) and the plan of reopening (May 11, 2020). By comparison, the reproductive number estimates based on the reported curve are described by the dashed curve.

**Figure 7:**
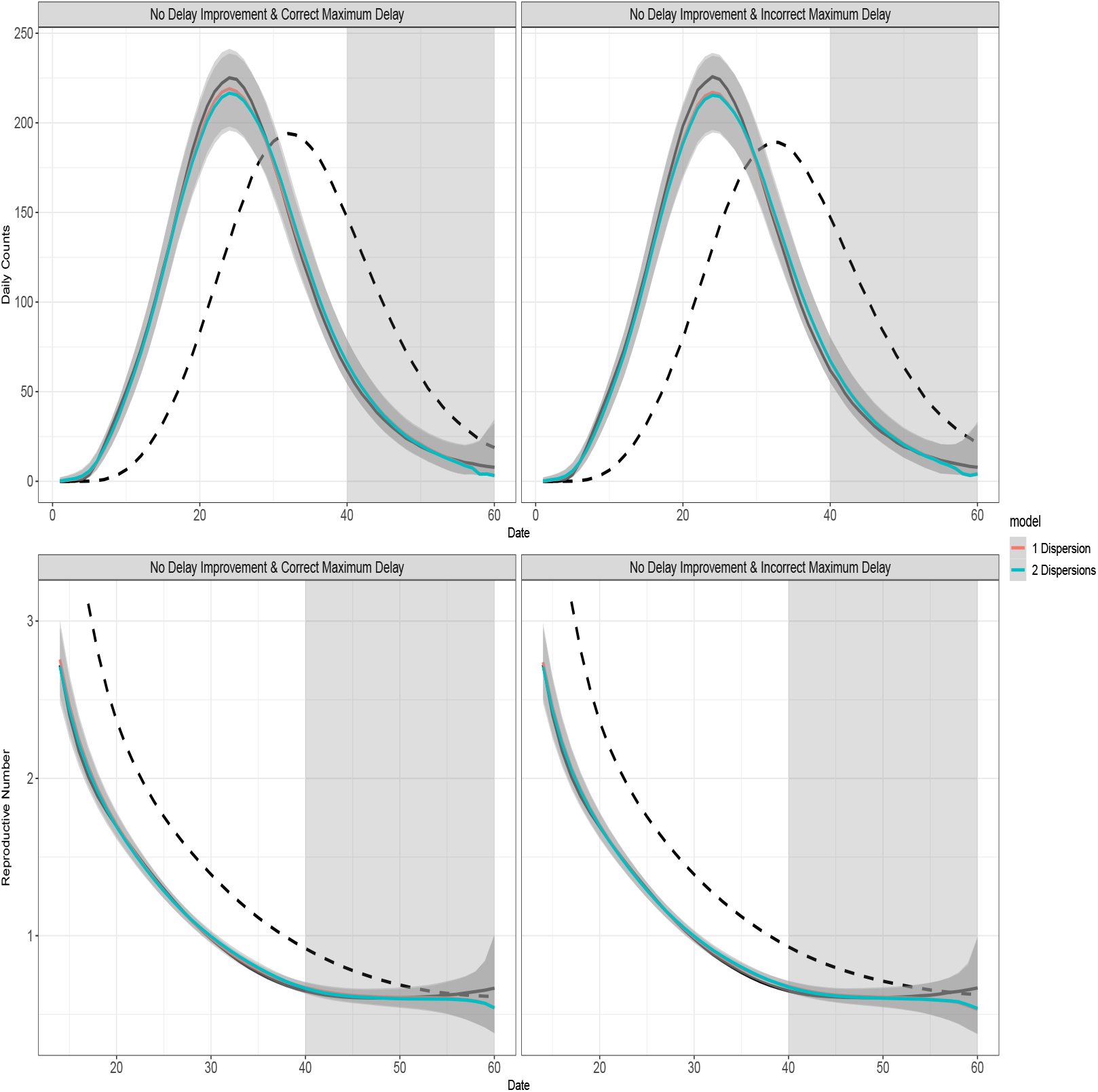
Impact of the maximum delay assumption for complete data when the reporting delay distribution was unchanged. For all graphs: the black solid curve corresponds to estimates based on the known epidemic curves and the black dashed curve corresponds to estimates based on the reported curves. The grey-shaded region superimposed on the curve depicts the 95% Bayesian credible interval and the grey-shade region on the right indicates the region of nowcasting. The colored curves represent different model choices. All values were averaged over 1000 simulated datasets.

**Figure 8:**
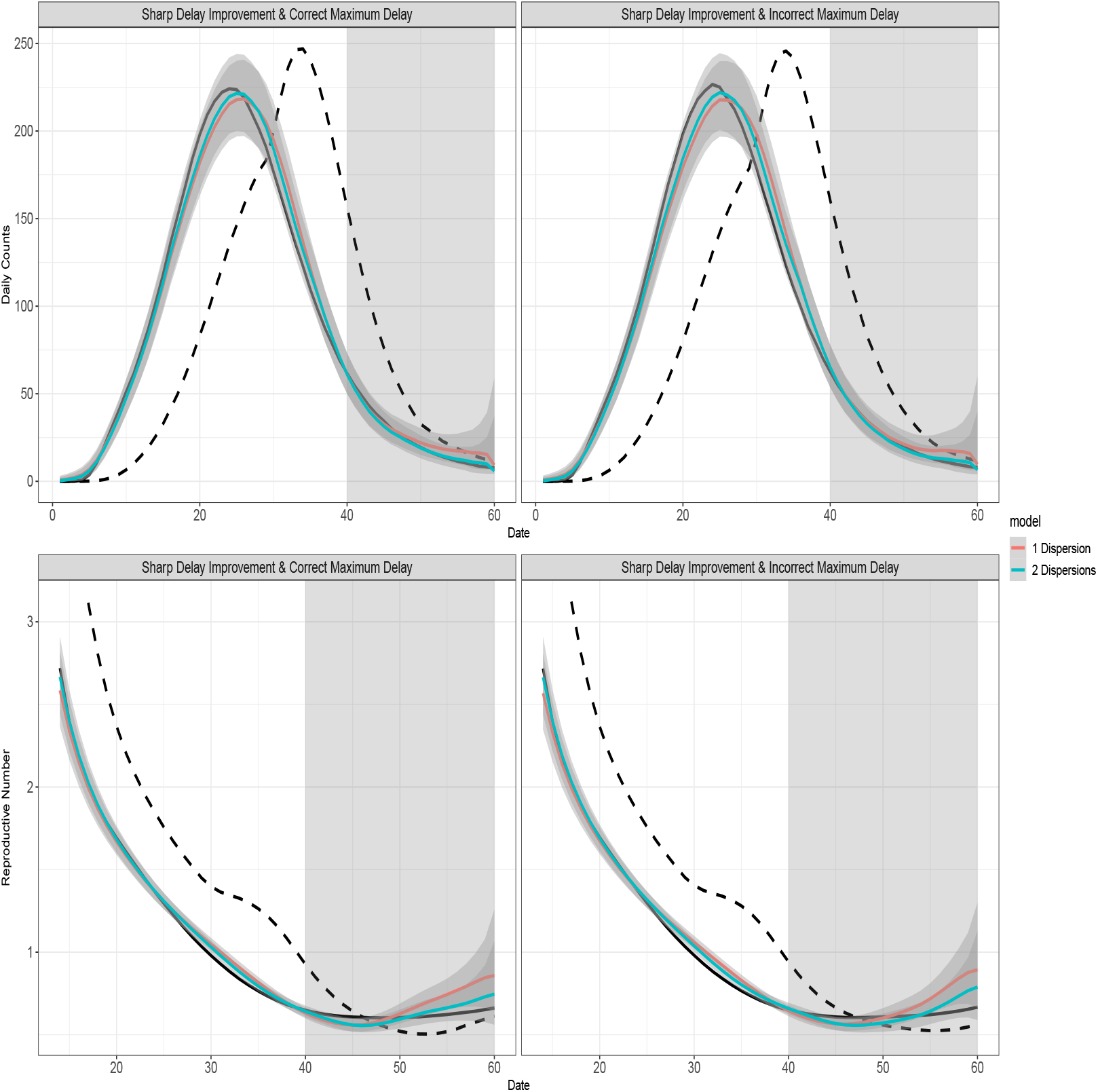
Impact of the maximum delay assumption for complete data when the reporting delay distribution was sharply improved. For all graphs: the black solid curve corresponds to estimates based on the known epidemic curves and the black dashed curve corresponds to estimates based on the reported curves. The grey-shaded region superimposed on the curve depicts the 95% Bayesian credible interval and the grey-shade region on the right indicates the region of nowcasting. The colored curves represent different model choices. All values were averaged over 1000 simulated datasets.

**Figure 9:**
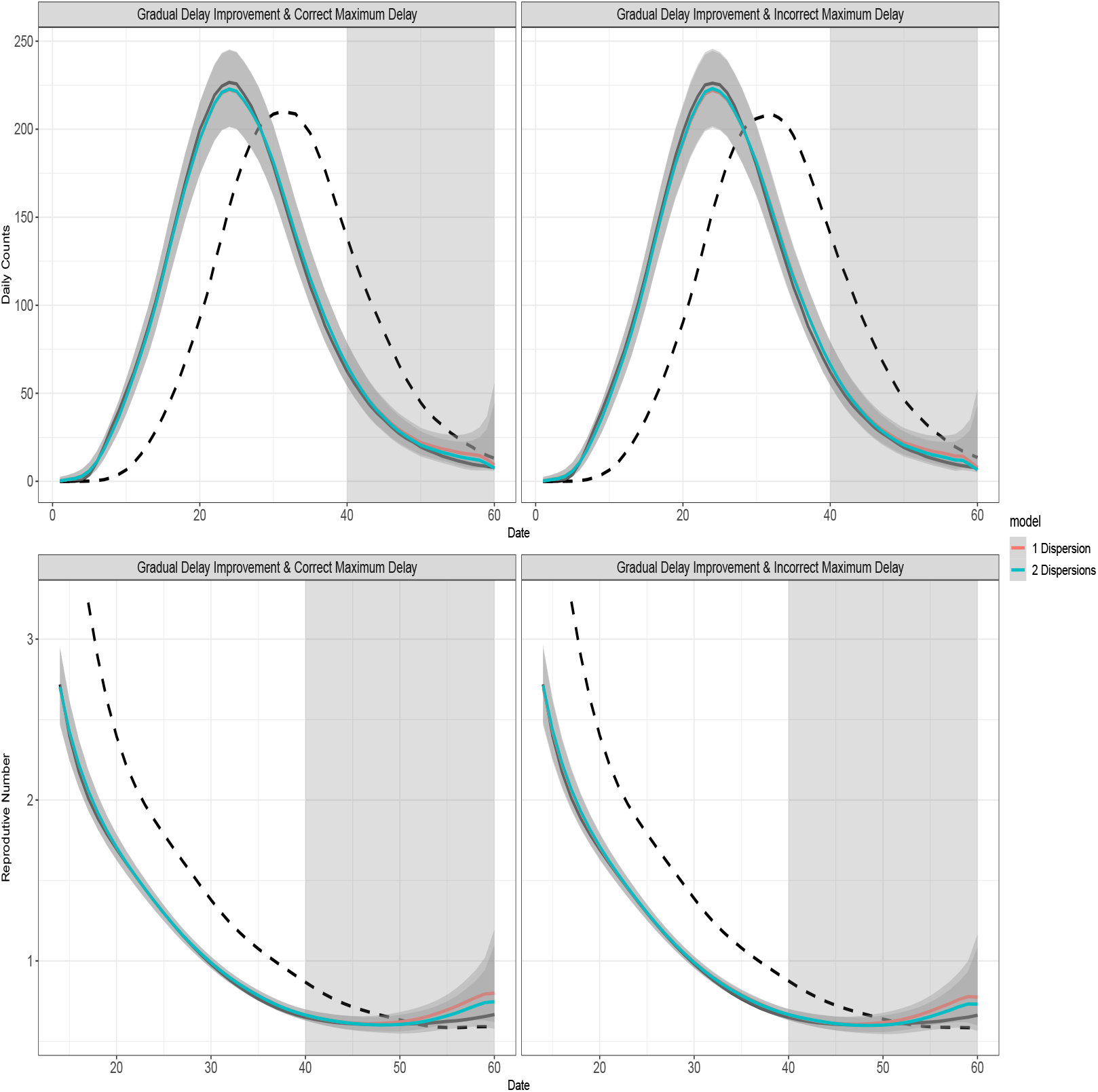
Impact of the maximum delay assumption for complete data when the reporting delay distribution was gradually improved. For all graphs: the black solid curve corresponds to estimates based on the known epidemic curves and the black dashed curve corresponds to estimates based on the reported curves. The grey-shaded region superimposed on the curve depicts the 95% Bayesian credible interval and the grey-shade region on the right indicates the region of nowcasting. The colored curves represent different model choices. All values were averaged over 1000 simulated datasets.

Our estimates under the scenario where the reporting delay distribution improved sharply at day 30 during the epidemic wave (Fig 2 C-D) was comparatively worse than other two scenarios (i.e., when the reporting delay distribution was either not improved or gradually improved), as manifested by the underestimation of the reporting delays between day 15 and day 40 during the epidemic wave. The underestimation was mainly due to the overlap of the two reporting delay distributions for cases reported from day 30 to day 50 (most of whom had symptom onsets from day 15 to day 40). Our model struggled to separately estimate the two distributions during this period because it is built on case reporting dates rather than symptom onset dates. We also used both the Eq (1) and Eq (2) for imputation and estimation. The two models performed similarly when the reporting delay distribution was unchanged or gradually improved. However, Eq (2) did result in a slightly better fit than Eq (1) when the reporting delay distribution sharply improved, likely due to having two dispersion parameters.

Table 1 lists the coverage rate of 95% Bayesian credible interval and the RMSE for our estimates. The average coverage rate of our epidemic curve estimates was 0.83 when there was an abrupt improvement for the reporting delay distribution and was 0.91 when there was gradual or no change in the reporting delay distribution. The average coverage rates of the reproductive number estimates was slightly lower than the average coverage rates of the epidemic curve estimates in general, likely due to the additional error brought by *EpiEstim* [21]. Compared to Eq (1), Eq (2) had higher coverage rates and RMSE of the epidemic curve estimates when the reporting delay distributions sharply improved at day 30 (coverage rate: from 0.83 to 0.90; RMSE: from 9.69 to 8.04). The gain of using Eq (2) was even larger for the reproductive number estimates in this case: the coverage rate increased from 0.58 to 0.74 and the RMSE decreased from 0.07 to 0.05. For the other two scenarios, Eq (2) was comparable to Eq (1). Overall, the Bayesian credible interval was tight (indicated by the small RMSE) with acceptable coverage rate (around 0.9) given appropriate model choice, when the line list data was complete for the epidemic wave.

**Table 1:** Performance measures for complete data. The results were averaged over all simulated datasets and dates for both the epidemic curve (Curve) and the reproductive numbers (*R*_*t*_). The results format: coverage rate (RMSE). Model 1 refers to the model in Eq (1) and model 2 refers to the model in Eq (2).

| Improvement | Maximum Delay | Model 1 |  | Model 2 |  |
| --- | --- | --- | --- | --- | --- |
| | | Curve | $R_t$ | Curve | $R_t$ |
| No | Correct | 0.92<br>(7.48) | 0.89<br>(0.05) | 0.92<br>(7.72) | 0.87<br>(0.05) |
|  | Incorrect | 0.91<br>(7.94) | 0.85<br>(0.05) | 0.90<br>(8.13) | 0.85<br>(0.05) |
| Sharp | Correct | 0.83<br>(9.69) | 0.58<br>(0.07) | 0.90<br>(8.04) | 0.74<br>(0.05) |
|  | Incorrect | 0.79<br>(10.59) | 0.59<br>(0.07) | 0.88<br>(8.66) | 0.70<br>(0.05) |
| Gradual | Correct | 0.90<br>(7.69) | 0.85<br>(0.05) | 0.92<br>(7.23) | 0.89<br>(0.05) |
|  | Incorrect | 0.89<br>(7.87) | 0.84<br>(0.05) | 0.92<br>(7.37) | 0.88<br>(0.05) |

We also checked the coverage rate and RMSE of the case count estimates on each day based on symptom onset dates, in order to evaluate the performance of our model from a temporal perspective (Fig 10 and Fig 11). Overall, the coverage rate was negatively correlated with the RMSE, consistent with of our other results. The coverage rate of our estimate was consistently over 0.9 except the last several days when the reporting delay distribution did not sharply improve during the epidemic wave. There are three reasons for this. First, back-calculation performed well in our model and led to accurate estimates of daily case counts. Second, nowcasting led to good estimates until the last few days, suggesting our nowcasting framework is valid. Third, nowcasting led to poor estimates for the last several days as the case counts of those days suffered most from the right truncation issue, i.e., most of those case counts were to be reported after the final reporting date (day 60) of the line-list data and thus unavailable for analysis. Therefore, the poor nowcasting performance was excusable given the right truncation issue was the worst for the last several days.

**Figure 10:**
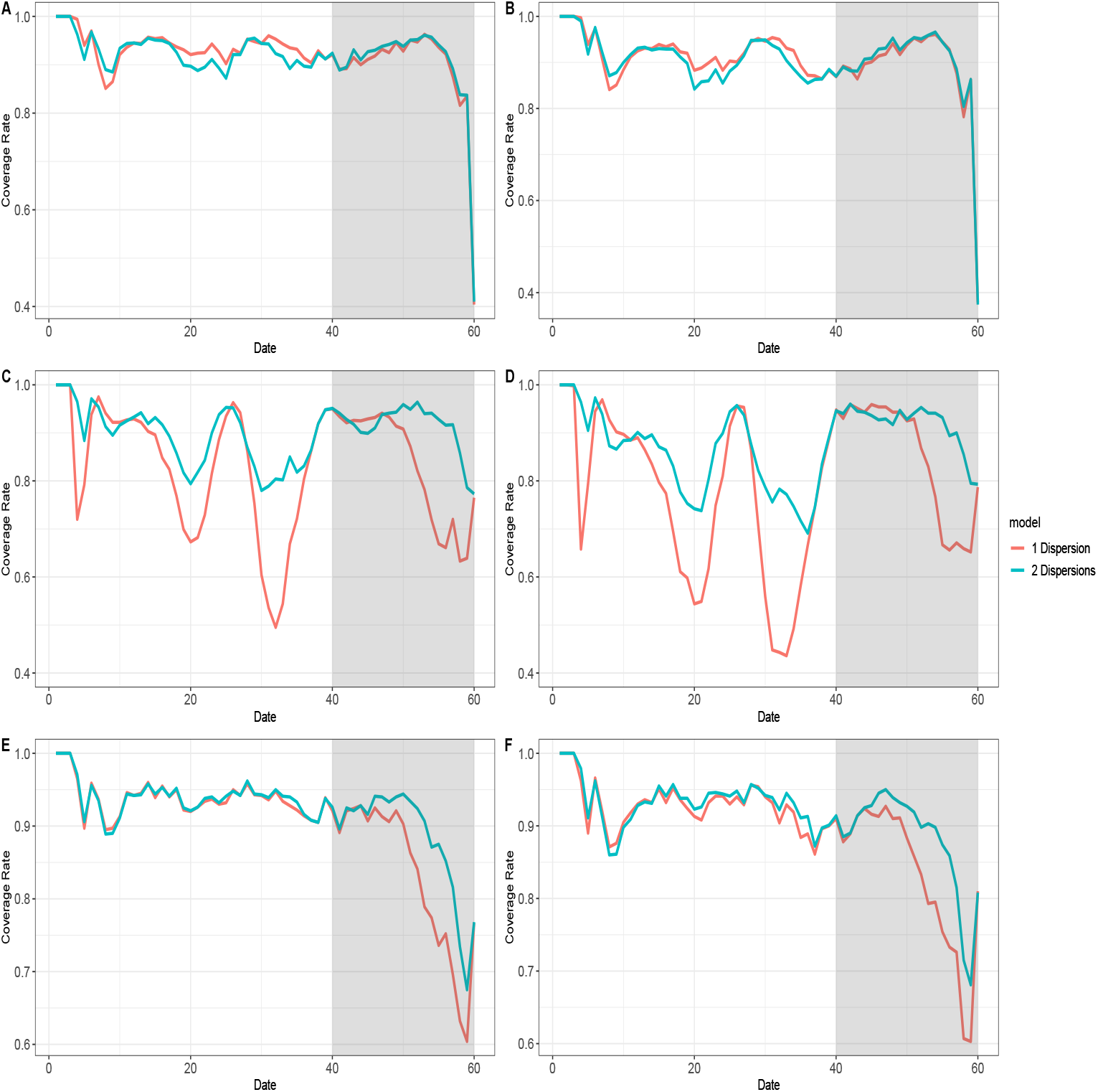
Coverage rates of all estimated daily counts of symptom onset cases for complete data. For all graphs: The colored curves represent different model choices and the grey-shaded region indicates the nowcasting region. The coverage rates were calculated based on 1000 simulated datasets. A: The coverage rates given the reporting delay distribution was unchanged and *l* was correct. B: The coverage rates given the reporting delay distribution was unchanged and *l* was incorrect. C: The coverage rates given the reporting delay distribution was sharply improved and *l* was correct. D: The coverage rates given the reporting delay distribution was sharply improved and *l* was incorrect. E: The coverage rates given the reporting delay distribution was gradually improved and *l* was correct. F: The coverage rates given the reporting delay distribution was gradually improved and *l* was incorrect.

**Figure 11:**
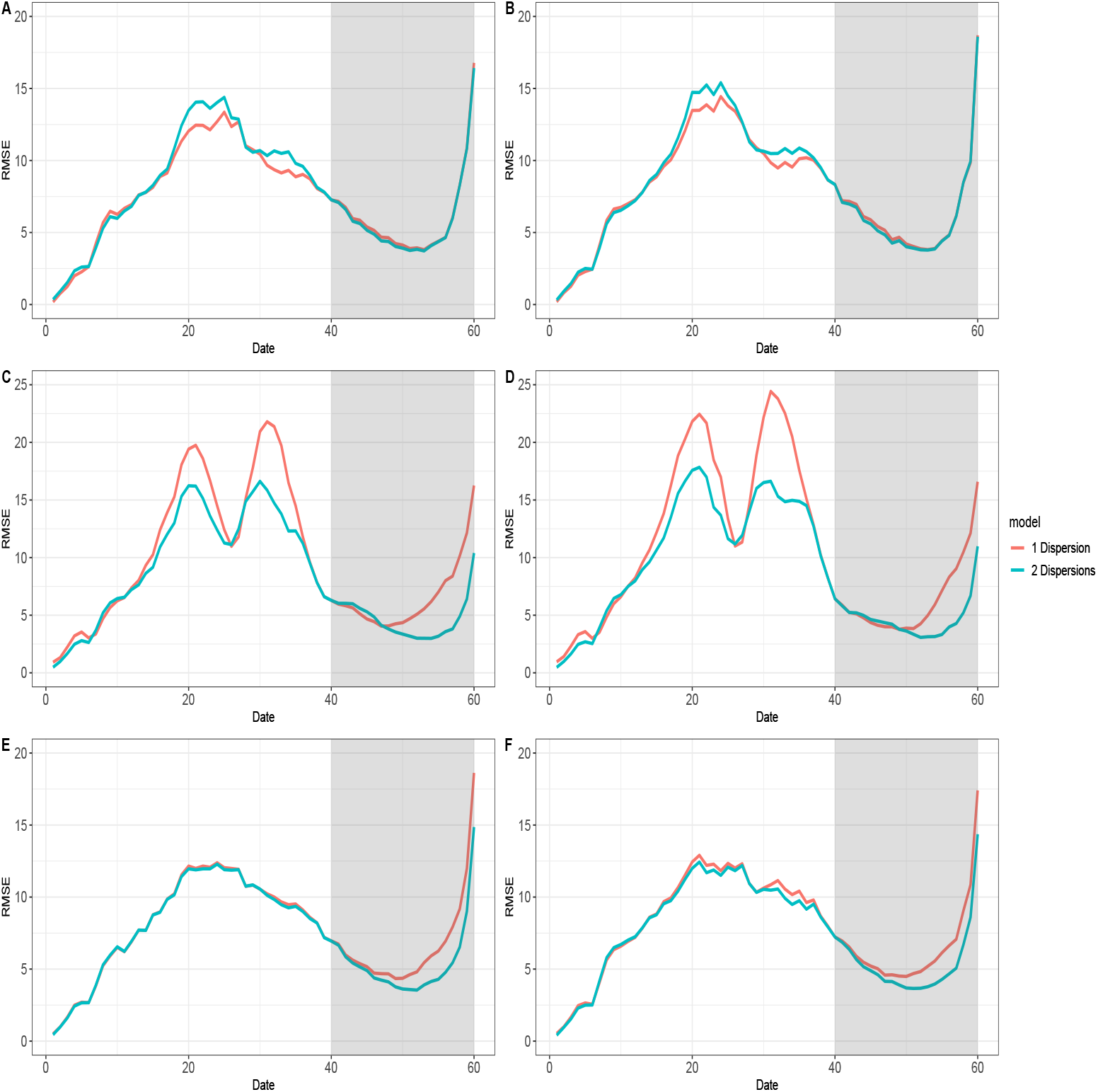
RMSE of all estimated daily counts of symptom onset cases for complete data. For all graphs: The colored curves represent different model choices and the grey-shaded region indicates the nowcasting region. The RMSE were calculated based on 1000 simulated datasets. A: The RMSE given the reporting delay distribution was unchanged and *l* was correct. B: The RMSE given the reporting delay distribution was unchanged and *l* was incorrect. C: The RMSE given the reporting delay distribution was sharply improved and *l* was correct. D: The RMSE given the reporting delay distribution was sharply improved and *l* was incorrect. E: The RMSE given the reporting delay distribution was gradually improved and *l* was correct. F: The RMSE given the reporting delay distribution was gradually improved and *l* was incorrect.

For delayed surveillance initiation, we assume four different starting dates for the line list data: day 11, day 21, day 31, and day 41 (see appendix for more details). To enhance comparability of the results based on the line-list data with different starting dates, we only used the Eq (1) for estimation. In general, we estimate the epidemic curve well from the starting date onward (Fig 3). For reproductive number estimation, the estimates become reliable *τ* + 1 days after the starting date, since *EpiEstim* needs at least *τ* + 1 days’ observations to produce unbiased estimates. For example, if the starting date is day 11 and *τ* = 6 one should expect the epidemic curve and reproductive number estimates to converge to their benchmarks from day 11 and day 18 respectively. In general, estimation accuracy decreases with longer delays (Table 2). For individual daily case counts, the coverage rate (Fig 12) and the RMSE (Fig 13) were acceptable after the starting date, and the impact on the nowcasted case counts was minimal. We still observe that the estimated epidemic curve and reproductive numbers were far better than the reported curve and its associated reproductive numbers, unless there was a severe loss of early reporting (eg. if the starting date was day 31 or 41).

**Table 2:** Performance measures for data with no early report. The results were averaged over all simulated datasets and dates for both the epidemic curve (Curve) and the reproductive numbers (*R*_*t*_). The line-list data could start on day 11 (Data 1), day 21 (Data 2), day 31 (Data 3) or day 41 (Data 4). The results format: coverage rate (RMSE).

| Improvement | Maximum Delay | Data 1 |  | Data 2 |  | Data 3 |  | Data 4 |  |
| --- | --- | --- | --- | --- | --- | --- | --- | --- | --- |
| | | Curve | $R_t$ | Curve | $R_t$ | Curve | $R_t$ | Curve | $R_t$ |
| No | Correct | 0.91<br>(7.79) | 0.85<br>(0.07) | 0.71<br>(13.61) | 0.59<br>(0.30) | 0.52<br>(37.97) | 0.41<br>(0.58) | 0.36<br>(67.50) | 0.20<br>(0.67) |
|  | Incorrect | 0.90<br>(8.48) | 0.82<br>(0.07) | 0.70<br>(14.04) | 0.56<br>(0.28) | 0.52<br>(37.90) | 0.40<br>(0.51) | 0.36<br>(66.82) | 0.20<br>(0.57) |
| Sharp | Correct | 0.83<br>(10.16) | 0.60<br>(0.08) | 0.62<br>(16.08) | 0.29<br>(0.25) | 0.48<br>(39.98) | 0.30<br>(0.56) | 0.35<br>(71.52) | 0.14<br>(0.60) |
|  | Incorrect | 0.79<br>(10.98) | 0.60<br>(0.08) | 0.61<br>(16.73) | 0.34<br>(0.24) | 0.46<br>(40.20) | 0.33<br>(0.50) | 0.35<br>(71.04) | 0.18<br>(0.52) |
| Gradual | Correct | 0.89<br>(8.29) | 0.82<br>(0.07) | 0.68<br>(14.55) | 0.51<br>(0.29) | 0.49<br>(42.01) | 0.33<br>(0.61) | 0.34<br>(70.82) | 0.13<br>(0.77) |
|  | Incorrect | 0.89<br>(8.19) | 0.82<br>(0.06) | 0.68<br>(14.65) | 0.51<br>(0.28) | 0.49<br>(41.69) | 0.33<br>(0.55) | 0.33<br>(70.39) | 0.14<br>(0.69) |

**Figure 12:**
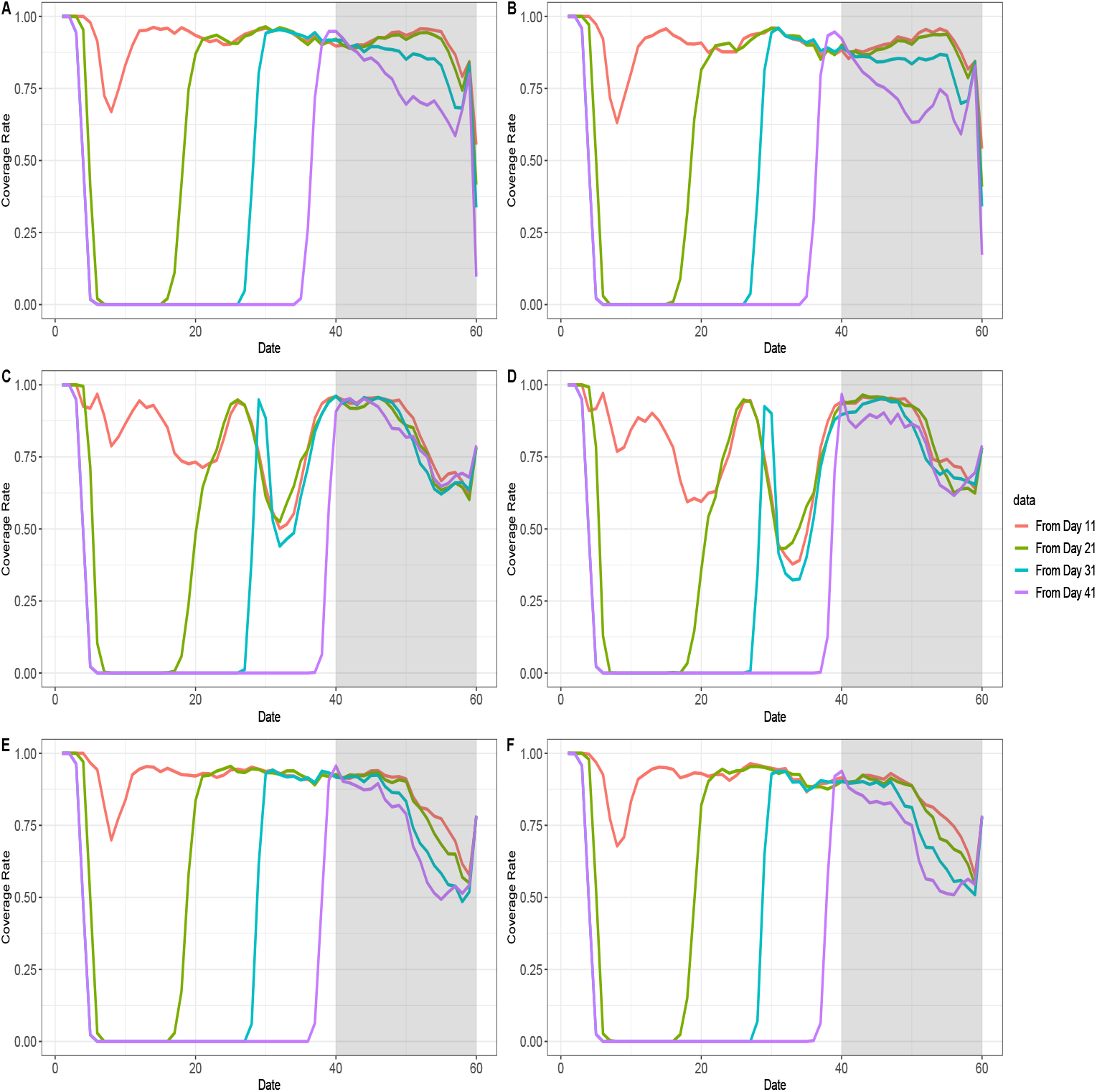
Coverage rates of all estimated daily counts of symptom onset cases for data with no early report. For all graphs: The colored curves represent different starting dates for line-list data and the grey-shaded region indicates the nowcasting region. The coverage rates were calculated based on 1000 simulated datasets. A: The coverage rates given the reporting delay distribution was unchanged and *l* was correct. B: The coverage rates given the reporting delay distribution was unchanged and *l* was incorrect. C: The coverage rates given the reporting delay distribution was sharply improved and *l* was correct. D: The coverage rates given the reporting delay distribution was sharply improved and *l* was incorrect. E: The coverage rates given the reporting delay distribution was gradually improved and *l* was correct. F: The coverage rates given the reporting delay distribution was gradually improved and *l* was incorrect.

**Figure 13:**
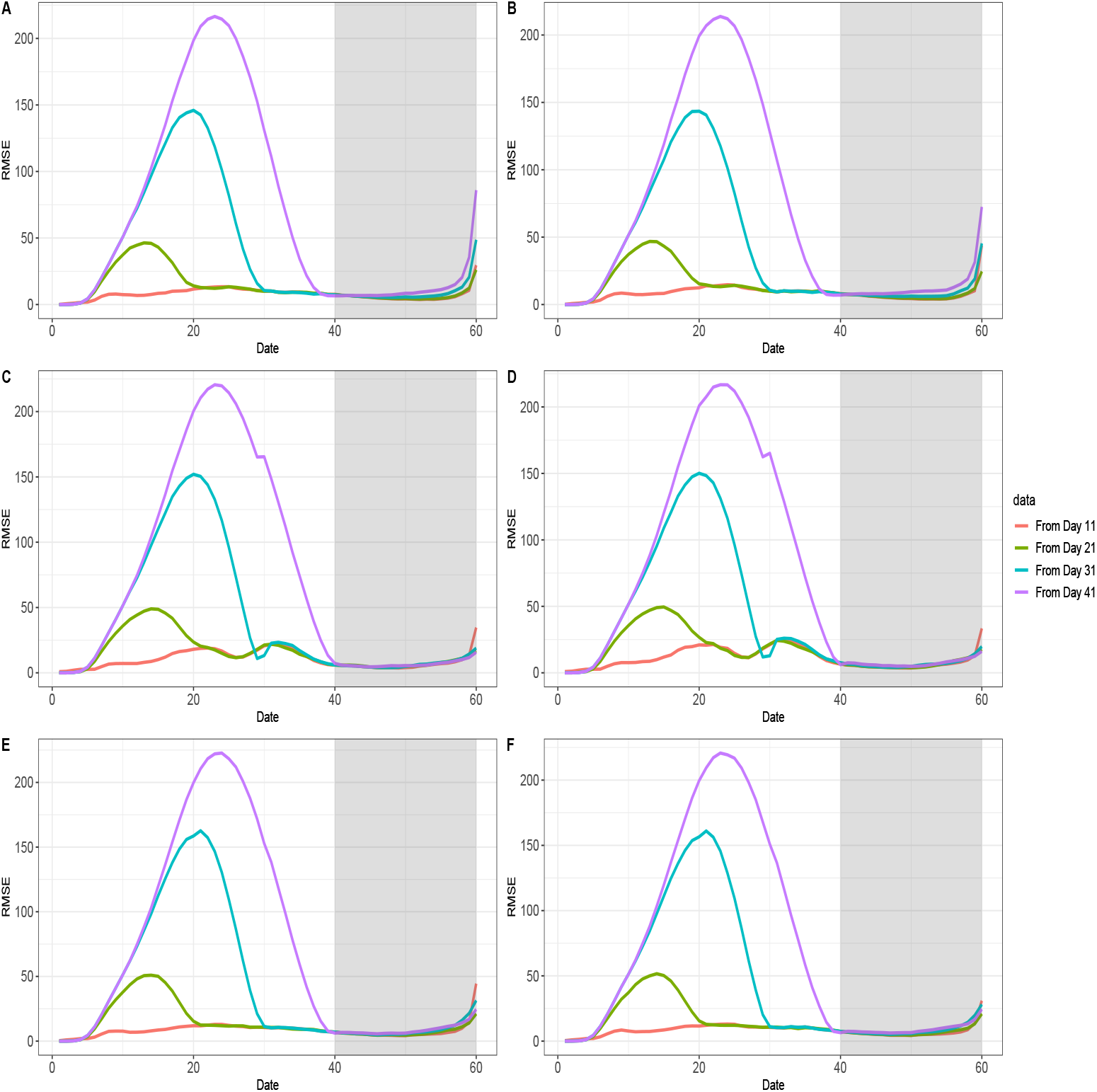
RMSE of all estimated daily counts of symptom onset cases for data with no early report. For all graphs: The colored curves represent different starting dates for line-list data and the grey-shaded region indicates the nowcasting region. The RMSE were calculated based on 1000 simulated datasets. A: The RMSE given the reporting delay distribution was unchanged and *l* was correct. B: The RMSE given the reporting delay distribution was unchanged and *l* was incorrect. C: The RMSE given the reporting delay distribution was sharply improved and *l* was correct. D: The RMSE given the reporting delay distribution was sharply improved and *l* was incorrect. E: The RMSE given the reporting delay distribution was gradually improved and *l* was correct. F: The RMSE given the reporting delay distribution was gradually improved and *l* was incorrect.

### 3.2 Simulation results: real time estimation

We chose day 28 (before the peak) or day 38 (after the peak) as the final reporting dates for the line-list data. As in the previous section, we only used Eq (1) for estimation to ensure comparability of the results. When using 28 days of data, we consistently underestimated the epidemic curve and the reproductive numbers (Fig 4). The average coverage rates were low (epidemic curve: 0.52, reproductive number: 0.37) and the RMSE were large (epidemic curve: 34.45, reproductive number: 0.15). By comparison, the average coverage rates were much higher if the final reporting date was day 38 (epidemic curve: 0.81, reproductive number: 0.74), and in this case the RMSE were much lower (epidemic curve: 16.67, reproductive numbers: 0.07) (Table 3).

**Table 3:** Performance measures for an ongoing epidemic wave. The results were averaged over all simulated datasets and dates for both the epidemic curve (Curve) and the reproductive numbers (*R*_*t*_). The line-list data could end on day 28 (Data 1) or day 38 (Data 2). The results format: coverage rate (RMSE).

| Improvement | Maximum Delay | Data 1 |  | Data 2 |  |
| --- | --- | --- | --- | --- | --- |
| | | Curve | $R_t$ | Curve | $R_t$ |
| No | Correct | 0.50<br>(37.74) | 0.31<br>(0.17) | 0.82<br>(15.69) | 0.79<br>(0.06) |
|  | Incorrect | 0.45<br>(40.14) | 0.31<br>(0.17) | 0.74<br>(17.83) | 0.78<br>(0.06) |
| Sharp | Correct | 0.50<br>(37.67) | 0.31<br>(0.17) | 0.67<br>(23.60) | 0.56<br>(0.10) |
|  | Incorrect | 0.46<br>(40.20) | 0.31<br>(0.17) | 0.62<br>(24.68) | 0.54<br>(0.11) |
| Gradual | Correct | 0.55<br>(27.95) | 0.48<br>(0.12) | 0.93<br>(10.73) | 0.86<br>(0.05) |
|  | Incorrect | 0.52<br>(29.43) | 0.52<br>(0.11) | 0.92<br>(11.19) | 0.86<br>(0.05) |

Interestingly, our model had the best performance when there was gradual improvement in the reporting delay distribution, especially if the final reporting date was day 38 for the line list data. In this case, the coverage rates and RMSE of the estimates were very close to those for complete data, and the coverage rates were consistently around 0.9 for all individual daily case counts (Fig 14, Fig 15). In the other two scenarios, the coverage rates and RMSE were much worse and very unstable. This is because our model approximated the gradually improving reporting delay distribution well as it was built on the small reporting periods, which could be perceived as smoothing windows and lead to good local estimates. This is also because most part of the epidemic curve and reproductive number estimation was done by nowcasting (Fig 4), which benefits from gradually improved reporting delays. When there was no improvement in the reporting delay distribution, underestimation is worse due to more extreme right truncation. When there was a sharp improvement for the reporting delay distribution, we observed an erratic sudden jump of the daily count estimates, which likely resulted from nowcasted case counts being overweighted as reporting delays tended to be underestimated, a pattern that had been observed for the complete data.

**Figure 14:**
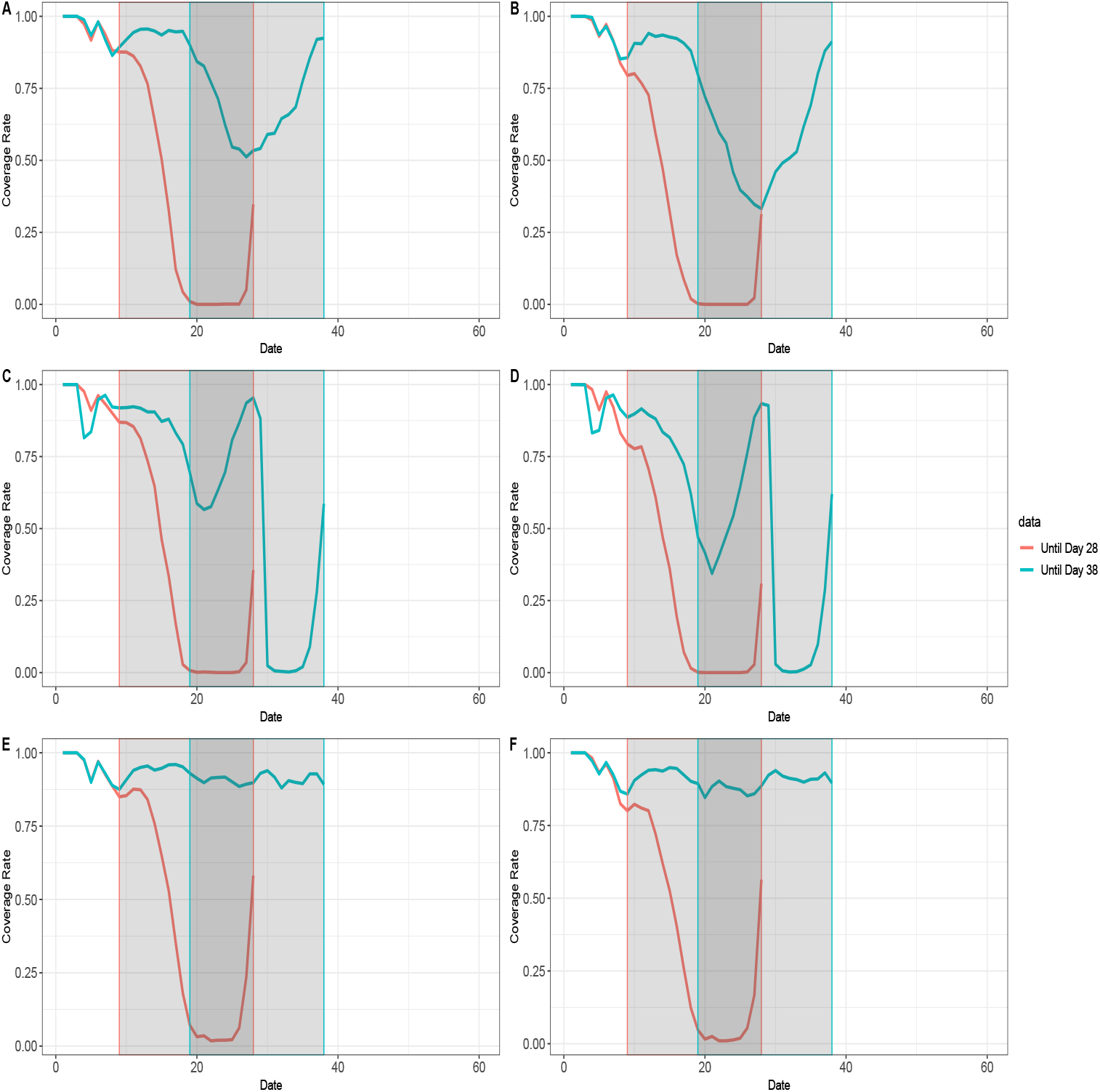
Coverage rates of all estimated daily counts of symptom onset cases for an ongoing epidemic wave. For all graphs: The colored curves represent different ending dates for line-list data, and their nowcasting regions are displayed as the gray-shaded areas with boundary lines in their corresponding colors. The coverage rates were calculated based on 1000 simulated datasets. A: The coverage rates given the reporting delay distribution was unchanged and *l* was correct. B: The coverage rates given the reporting delay distribution was unchanged and *l* was incorrect. C: The coverage rates given the reporting delay distribution was sharply improved and *l* was correct. D: The coverage rates given the reporting delay distribution was sharply improved and *l* was incorrect. E: The coverage rates given the reporting delay distribution was gradually improved and *l* was correct. F: The coverage rates given the reporting delay distribution was gradually improved and *l* was incorrect.

**Figure 15:**
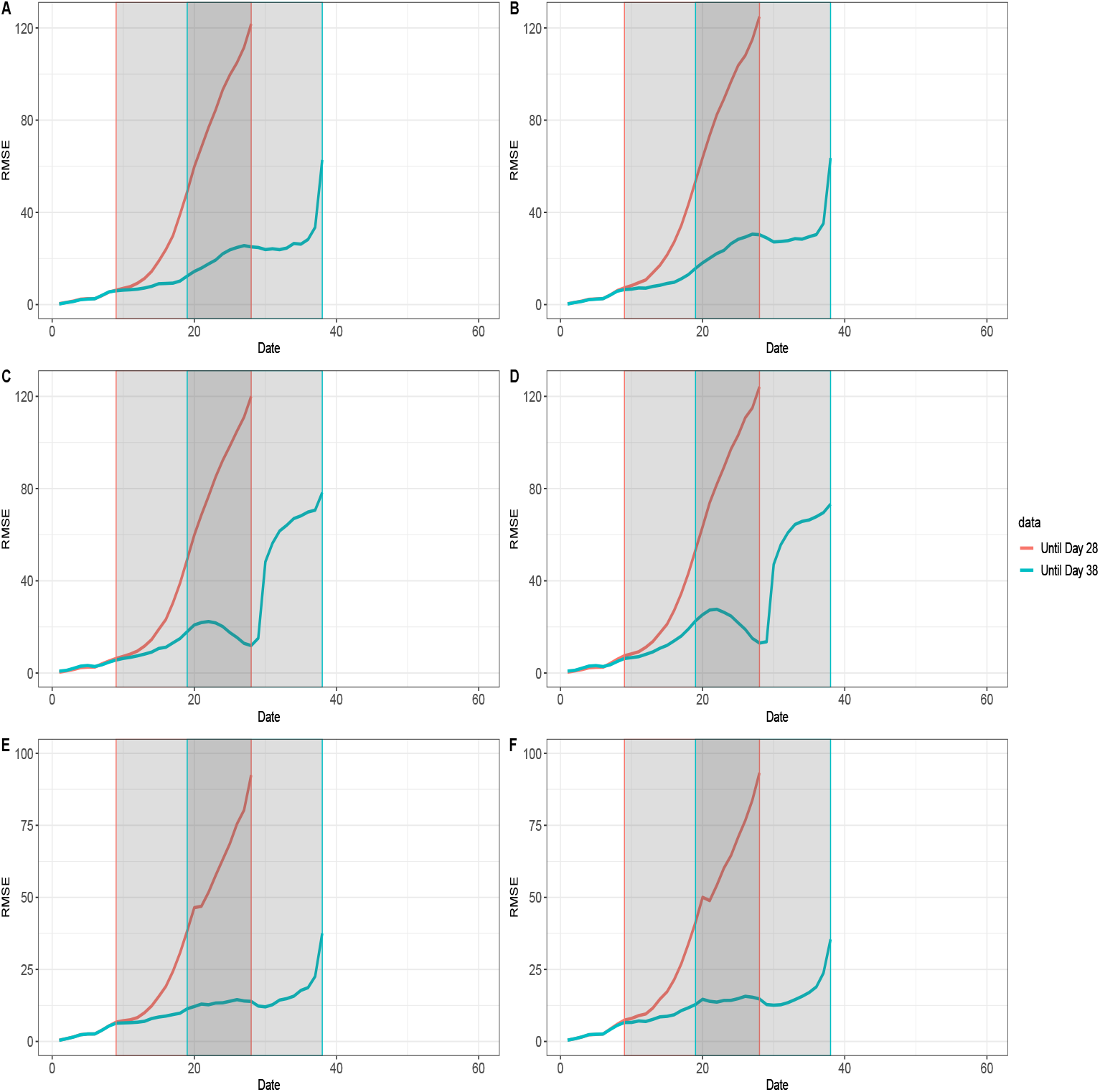
RMSE of all estimated daily counts of symptom onset cases for an ongoing epidemic wave. For all graphs: The colored curves represent different ending dates for line-list data, and their nowcasting regions are displayed as the gray-shaded areas with boundary lines in their corresponding colors. The RMSE were calculated based on 1000 simulated datasets. A: The RMSE given the reporting delay distribution was unchanged and *l* was correct. B: The RMSE given the reporting delay distribution was unchanged and *l* was incorrect. C: The RMSE given the reporting delay distribution was sharply improved and *l* was correct. D: The RMSE given the reporting delay distribution was sharply improved and *l* was incorrect. E: The RMSE given the reporting delay distribution was gradually improved and *l* was correct. F: The RMSE given the reporting delay distribution was gradually improved and *l* was incorrect.

### 3.3 COVID-19 in Massachusetts

We estimated the epidemic curve based on the COVID-19 line list data in Massachusetts and compared it with the reported curve (Fig 5). The estimated epidemic curve was much smoother than the reported curve, which indicates that most of the fluctuations were artificial. Based on the epidemic curve, we estimate that the COVID-19 outbreak started in early March in Massachusetts, and the daily count of cases showing symptoms began to decline around mid April with a slight increase around May 10. We estimated time-varying reproductive numbers assuming the distribution of serial interval is Gamma(4.29, 1.18) and *τ* = 6. The reproductive number estimates decreased from above 2 at the start of the outbreak to below 1.5 when lockdown that was enacted on March 17 (Fig 6). The stay-at-home order (issued by the Governor on March 23) further reduced the reproductive number below 1. The reopening plan was unveiled on May 11, at which point the reproductive number began to increase again. The reproductive number estimates based on the estimated epidemic curve corresponded more closely to these control measures than those based on the reported curve.

## 4 Discussion

Reproductive numbers are urgently needed for monitoring the progression of the COVID-19 pandemic, and they should be estimated based on reliable epidemic curve estimates, rather than the reported curve, to ensure minimal loss of the epidemiological signal. We introduce a Bayesian approach to estimate the epidemic curve and time-varying reproductive numbers from line list data. This approach has two unique advantages over other similar approaches. First, it is built on line list data which contains individual reporting delays that allow the estimation of the reporting delay distribution to be data-oriented and time-dependent. Second, it integrates the tasks of estimation of the reporting delay distribution, imputation of the reporting delay as well as estimation of the epidemic curve and reproductive numbers into one Bayesian framework, making those three tasks interdependent. As a result, our approach more accurately estimates uncertainty and is more efficient than other approaches that perform the three tasks independently. The results suggest the Bayesian approach is robust to unfavorable changes in data availability and misspecification of the reporting delay or the maximum delay assumption. Under typical assumptions, the Bayesian approach produces accurate estimates (low RMSE) and reliable inference (high coverage rate).

Unsurprisingly, the model performance does rely on data availability, and it will be inadequate based on insufficient data. For a single epidemic wave, our model estimates both the epidemic curve and reproductive numbers well if line list data is available for the whole epidemic wave, though one should be cautious about the model choice if the reporting system has significantly improved over time. If there are severe delays in initiating surveillance, our model will likely underestimate the case counts of the days prior to the starting date of surveillance, and the *R*_*t*_ estimates will eventually converge at a rate consistent with the serial interval. If estimation is performed in the midst of an outbreak, the Bayesian approach will underestimate the epidemic curve before the peak of the reported curve but performs substantially better after the peak. This suggests that, in the case of single epidemic wave, we need to wait until the peak of the reported curve has passed to ensure there is sufficient data for estimating the reproductive number using this approach. We stress that, if a line list data contains multiple epidemic waves, the Bayesian estimates are at least accurate for all except the last epidemic wave. To safely estimate the last epidemic wave, one still needs to wait until the majority of its cases are reported.

The model is sensitive to sharp changes in the reporting delay distribution. If the reporting delay distribution remains unchanged or changes gradually, our model generally performs well. Nowcasting performance is actually improved when the reporting delay distribution improves over time, due to shorter reporting delays. However, if there is a sharp improvement for the reporting delay distribution, our model will generate inaccurate estimates during the period when the two underlying reporting delay distributions overlap, resulting in underestimation of reporting delays. In this case, it would be beneficial to use Eq (2) to fit the reporting delay distribution instead. In general, we recommend using Eq (2) for the reporting delay distributions with changes and Eq (1) for those without changes.

Our model generates a posterior sample of time-varying reproductive number estimates, based on the epidemic curve estimates. We use *EpiEstim* to compute time-varying reproductive numbers, conditional on the maximum length and distribution of serial interval. We choose *EpiEstim* because it is more appropriate for real-time analysis and tracking of temporal changes (such as impact of a policy), compared to other alternatives [17]. We recommend using an integrated approach that includes both inference of the reporting delays and estimation of reproductive numbers, to incorporate all sources of uncertainty in modeling, since we are better able to estimate variability due to estimation from this multistage process. We note a few limitations of our approach that are inherited from the *EpiEstim* estimator. First, the maximum length of serial interval *s* and the sliding window size *τ* are subjective choices [21]. Second, it is possible to have negative serial intervals for COVID-19 which is currently not allowed by *EpiEstim* [25]. Third, it is most accurate to estimate reproductive numbers from the incidence curve rather than the epidemic curve for *EpiEstim* [21, 26]. However, infection events would be very hard, if not impossible, to observe for the current pandemic and thus strong parametric assumptions are likely needed [12, 14], which is beyond the scope of this paper. Fourth, reproductive number estimates will be less trustworthy if the fraction of infection observed is not constant over time [3, 20, 27]. For COVID-19, this is likely the case considering the evolution of testing and the significant proportion of asymptomatic transmission [28], requiring further adjustment of the data.

Empirically, there are some important issues to consider in properly implementing our method. First, our model is region-specific, i.e., one need to fit our model to line list data of a single region to avoid systematic differences between regions. The region is defined such that each region is deemed to have its own reporting system (and thus its unique reporting delay distribution). For example, if the reporting system differs at the county level, we should use line list data of each county (rather than each state) for our model. Second, the reporting period in our model needs to be carefully and properly defined, as our model is essentially a moving-window smoothing method. As with most other moving-window smoothing methods, the model performance depends on the moving-window size, which in our case is the reporting period size [17]. The moving-window size is known for its pivotal role in the bias-variance trade-off and thus should be neither too small nor too large for estimating the reporting delay distribution [5]. Third, our model cannot handle negative reporting delays which are possible for the current COVID-19 pandemic due to contact tracing, though our assumption of non-negative reporting delays is consistent with the literature [3, 14].

Overall, we provide an useful tool to estimate timely reproductive number estimates based on a Bayesian approach that integrates reporting delay imputation, back-calculation and nowcasting, all of which are interdependent and critical for reproductive number estimation. Our approach is robust to reasonable deviations from the model assumptions. Most importantly, it is more epidemiological meaningful than estimates based on the reported curve and thus a better option for surveillance of the COVID-19 pandemic.

## Data Availability

Code and Output is available on github

https://github.com/tenglongli/backandnow

## Supporting information

