## Appendix for "Bayesian back-calculation and nowcasting for line list data during the COVID-19 pandemic"

November 23, 2020

#### 1 Simulation design

We simulated epidemics similar to Covid-19. We assumed the local epidemic started with 100 initial cases and do not allow for imported cases in the simulation. The time-varying reproductive number $R_t$ was set based on literature [1, 2], to reflect the covid epidemic at multiple stages such as initial outbreak, lockdown as well as lift of lockdown (Fig 1). The distribution of generation interval was gamma distribution with shape and rate equal to 4.29 and 1.18, respectively, consistent with the literature [3]. The distribution of incubation period was lognormal distribution with mean and standard deviation of 1.621 and 0.418 respectively [4, 5]. We considered three factors in the simulation design, namely the reporting delay distribution, the maximum delay assumption and the data availability. In total, we had 18 different simulation scenarios ($3 \times 3 \times 2$).

The reporting delay distribution could be one of the following three scenarios:

1. No improvement: The reporting delay distribution was negative binomial distribution with $\mu = 9$ and $r = 3$ and it did not change.
2. Sharp improvement: The reporting delay distribution was negative binomial distribution with $\mu = 9$ and $r = 3$ before day 30 and was

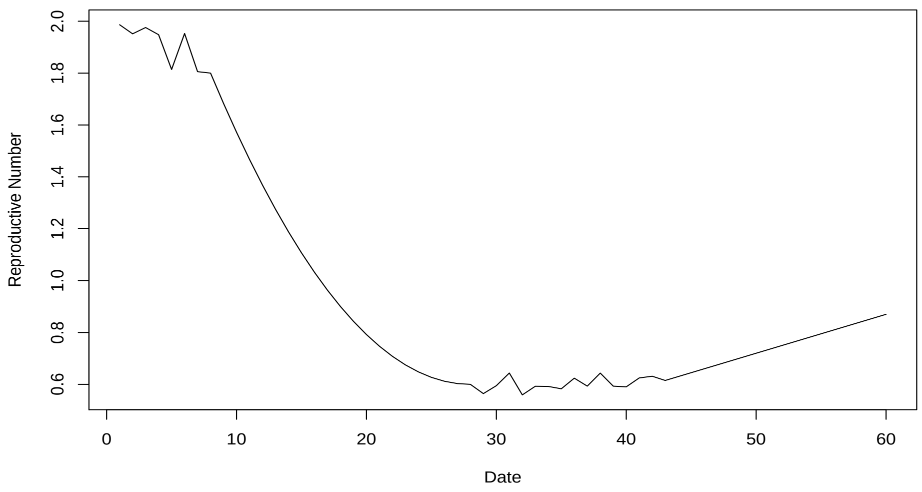


Figure 1: **The underlying time-varying reproductive numbers for the simulation.** This graph illustrates the time-varying reproductive numbers we used in the simulation. Notably, they corresponded to infection events from day 1 to day 60.

negative binomial distribution with $\mu = 4$ and $r = 3$ after day 30.

3. Gradual improvement: The reporting delay distribution was negative binomial distribution with $r$ fixed at 3. $\mu$ was set to 9 initially, then it got one day smaller every ten days from day 11 and finally $\mu = 4$ from day 51.

The maximum delay assumption could be one of the following two scenarios:

1. Correct maximum delay: $l$ was assumed to be 20 for our Bayesian algorithm and it was actually 20 in the simulation.

2. Incorrect maximum delay: $l$ was assumed to be 20 for our Bayesian algorithm and it was actually 25 in the simulation.

The data availability could be one of the following three scenarios:

1. Complete data: The line list data covered all cases reported from day 1 to day 60.

2. Delayed surveillance initiation: The line list data covered all cases reported from day $k$ to day 60. $k$ could be 11, 21, 31 or 41.

3. Real time estimation: The line list data covered all cases reported from day 1 to day $w$. $w$ could be either 28 or 38.

The simulation was done via branching process, i.e., we first drew the number of infectees from a Poisson distribution whose mean was defined by the number of infectors and corresponding reproductive number. We then obtained the infection dates, the symptom onset dates and the case reporting dates from the distributions of generation interval, incubation period and reporting delays respectively for those infectees. Finally, a line list data was created based on individual symptom onset dates and case reporting dates.

### 2 More discussion on delayed surveillance initiation

For delayed surveillance initiation, we assume four different starting dates for the line list data: day 11, day 21, day 31, and day 41. Cases reported earlier than the starting dates were not included in the line-list data for analysis. To enhance comparability of the results based on the line-list data with different starting dates, we only used the model 1 for estimation. In general, the starting date has a pretty straightforward and significant impact on the estimates. Specifically, we estimated the epidemic curve well from the starting date onward (Fig 3 in the main text). For reproductive number estimation, the date when the estimates began to be reliable was $\tau + 1$ days later than the starting date, since *EpiEstim* needed at least $\tau + 1$ days' observations to start in this case. For example, one should expect the epidemic curve and reproductive number estimates start to align well with their benchmarks from day 11 and day 18 respectively, if the starting date is day 11 and $\tau = 6$. It was clear that the estimates were worse if the line list data started later (Table 2 in the main text). For example, if the starting date was day 11 (data 1), the average coverage rate was 0.83 (RMSE: 10.16) when there was a sharp improvement for the reporting delay distribution and the average coverage rate was 0.9 (RMSE: 8.04) when there was not. This is very close to what we obtained using the complete data, and it suggested excluding the cases reported in the first 10 days should not matter in this case. However, if the starting date was day 21 (data 2), the coverage rate decreased to 0.62 (RMSE: 16.08) when there was a sharp improvement for the reporting delay distribution and to 0.7 (RMSE: 14.08) when there was not. For individual daily counts, we found again that the coverage rate (Supplementary figure 6) started to rise to an acceptable level after the starting date, and the lack of early reporting would cost the coverage rates of the nowcasted case counts slightly. The RMSE of back-calculation was unbearably large for dates prior to the starting date and dropped to an acceptable level after the starting date (Supplementary figure 7). We still observed that the estimated epidemic curve and reproductive numbers were far better than the reported curve and its associated reproductive numbers, unless there was a severe loss of early reporting (eg. if the starting date was day 31 or 41).

[2] Chong You, Yuhao Deng, Wenjie Hu, Jiarui Sun, Qiushi Lin, Feng Zhou, Cheng Heng Pang, Yuan Zhang, Zhengchao Chen, and Xiao-Hua Zhou. Estimation of the time-varying reproduction number of covid-19 outbreak in china. *International Journal of Hygiene and Environmental Health*, page 113555, 2020.

[3] Tapiwa Ganyani, Cécile Kremer, Dongxuan Chen, Andrea Torneri, Christel Faes, Jacco Wallinga, and Niel Hens. Authors' response: Esti-

mating the generation interval for COVID-19 based on symptom onset data. *Eurosurveillance*, 25(29):18–19, 2020.

[4] Stephen A. Lauer, Kyra H. Grantz, Qifang Bi, Forrest K. Jones, Qulu Zheng, Hannah R. Meredith, Andrew S. Azman, Nicholas G. Reich, and Justin Lessler. The incubation period of coronavirus disease 2019 (CoVID-19) from publicly reported confirmed cases: Estimation and application. *Annals of Internal Medicine*, 172(9):577–582, 2020.

[5] Natalie Linton, Tetsuro Kobayashi, Yichi Yang, Katsuma Hayashi, Andrei Akhmetzhanov, Sung-mok Jung, Baoyin Yuan, Ryo Kinoshita, and Hiroshi Nishiura. Incubation Period and Other Epidemiological Characteristics of 2019 Novel Coronavirus Infections with Right Truncation: A Statistical Analysis of Publicly Available Case Data. *Journal of Clinical Medicine*, 9(2):538, 2020.
